# Patients’ Ideas, Concerns, Expectations in Physiotherapy: A Questionnaire Study

**DOI:** 10.64898/2026.04.06.26350229

**Authors:** Ruchit Dani, Dhruvkumar Dave

## Abstract

Global healthcare is targeting patient-centred care, as it leads to better health outcomes and higher level of patient satisfaction. Patient-centred communication, is an important part of patient-centred care because it focuses on involving patients in their care. Recent surveys both nationally and globally have shown that patients are not involved enough in their own healthcare decisions. This problem is especially common among the elderly with chronic conditions. This study aimed to describe patient–healthcare professional interactions, expectations, and satisfaction in physiotherapy within an understudied context, thereby providing important, specific data on ICE dynamics and satisfaction in the specific setting. Cross-sectional study of participants in scheduled consultations was conducted. Two government physiotherapy centres, seven private physiotherapy centres and two trust centres with physiotherapy facilities in Gujarat, India. 232 patients (from various public and private physiotherapy clinics) participated in the study. Patients’ ideas, concerns, expectations (ICE) and satisfaction were explored. Almost 88% of patients reported their thoughts and explanations about their symptoms during the consultation. Most patients described not having any concerns about the diagnosis/treatment, and more than two-third of patients consulting PTs expected explanation for their symptoms. Almost 90% patients were satisfied with the consultation. The study revealed that while most patients conveyed their thoughts during consultations, very few expressed their concerns. Overall, patients were satisfied with their consultations.

## Introduction

While literature addresses ICE or expectations in musculoskeletal and chronic pain physiotherapy, direct trials isolating ICE-focused communication interventions and testing their impact on objective clinical outcomes (e.g., pain scores, function, return to work) are lacking. Evidence is therefore indirect and consensus is not established. Recent evidence has shown (indirectly) expectations, education, and adherence in musculoskeletal physiotherapy. In chronic nonspecific low back pain, patients reported high expectations of education from physiotherapists; clinicians mostly provided explanations of condition and prognosis, but education about causes, long term management, and tailored materials was often lacking ^1^. The author argued that aligning education with patient expectations is likely important for exercise adherence and outcomes, but this was a qualitative inference, not a tested causal effect ^1^.

A systematic review of high value physiotherapy for chronic pain identified that patient expectations and beliefs about treatment efficacy, the interrelationship with the physiotherapist, and the patient’s understanding of exercise benefits are major enablers or barriers to exercise adherence ^2^. Treatment expectations influenced adherence in patellofemoral pain, and perceived exercise efficacy predicted adherence in low back and other chronic pain conditions ^2^. The same review concluded that interventions to support high value care must address multifactorial barriers, including expectations, beliefs, and rapport, but the patient-facing interventions evaluated were mostly technology-based adherence tools, not explicit ICE style agenda elicitation ^2^. These findings suggest that exploring and aligning expectations and beliefs is plausibly important for adherence and hence, outcomes, but they do not demonstrate that formal ICE elicitation, as a technique, improves objective physiotherapy outcomes. What is known about ICE and satisfaction (mostly outside physiotherapy). In UK general practice, observational coding of 92 consultations found that most encounters contained at least one ICE/ICEE component; patient ideas were associated with a higher likelihood of patients being “very satisfied,” whereas concerns were associated with lower satisfaction ^3^.

Satisfaction gains were evident for patient-initiated ideas but not for GP-initiated ideas, suggesting that how ICE is elicited matters ^3^. A 2024 surgeon–patient video study found that explicit patient perspective (ICE) sequences occurred in < one-third of consultations; only 11% were initiated by surgeons ^4^. The authors highlighted that failing to incorporate patient perspectives risks lower satisfaction and poorer health outcomes, but the study was descriptive and did not quantify outcome changes ^4^. A 2024 primary care questionnaire study showed that patients and physicians diverge in their understanding of patient expectations: patients described a personal, longitudinal “journey,” whereas physicians framed expectations more in terms of system responsibilities ^5^. The authors concluded that better acceptance and meeting of expectations are important for patient-centred care but do not associate ICE absolutely to strong outcomes ^5^. Overall, there is consistent qualitative and observational evidence that ideas, concerns and expectations are essential to satisfaction and adherence, but the mechanism and dose–response of structured ICE techniques are uncertain.

Specific knowledge gaps for physiotherapy and satisfaction where evidence is clearly lacking are as follows: i) There is limited research on how structured ICE (Ideas, Concerns, and Expectations) interventions affect measurable results. Additionally, there is very little information describing patients ideas, concerns, expectations, and satisfaction during physiotherapy sessions, especially in different regions and cultures. (ii) No proven link between ICE and measureable outcomes in physio. Current physiotherapy research focuses on patient education, patient–therapist communication quality, and expectations, not on ICE per se, and does not test causal effects on standardised outcomes ^1 2^. (iii) Moreover, robust, context-specific data on patient satisfaction with physiotherapy settings are scarce, particularly regarding how ICE dynamics relate to overall satisfaction in non-Western countries ^1^. In reviews of high-value physiotherapy for chronic pain, reported that good rapport and patient perceived efficacy are facilitators, but patient satisfaction data are rarely reported or analysed ^2^. (iv) There is uncertainty about specific ICE components - like data suggest ideas and concerns might have opposite associations with satisfaction ^3^. It is unknown if similar patterns occur during physiotherapy, especially when pain, fear avoidance and long-term self management are important. (v) There is a significant knowledge gap because there has been little research on how ICE works and how satisfied patients are with physiotherapy in Indian healthcare. This is especially true for Gujarat, a region with people from many different economic backgrounds.

This study aimed to explore patient–professional interactions, expectations, and satisfaction in physiotherapy, thereby contributing basic insights into ICE dynamics and satisfaction in this specific understudied setting.

## Methods

### Study design

This cross-sectional study included patients who participated in scheduled consultations.

### Study setting

The study was conducted at 11 physiotherapy centres in Gujarat, India. These included two government centres, seven private centres, and two centres managed by trusts. The research was carried out over the period from January 20, 2023, to May 4, 2023. Out of 35 selected physiotherapy centres in Gujarat, India, all were invited to participate in the study. One of the researchers made invitations by telephone and in-person visits to the selected physiotherapy centres. Participating centres were purposively selected to facilitate initial data collection, prioritising sites with established relationships, reliable staff, and willingness to participate in the study. Although this approach was practical for this exploratory study and promoted participation, there may be potential selection bias, which is acknowledged as a limitation of generalisability. Six centres agreed to participate in the study, but one centre stopped participating after a few months because it did not have enough staff.

In Gujarat, socioeconomic status is in most cases high, and Gujarati is the most frequently used language. The population of one of the municipalities represented in the study has a higher educational level than that in most other areas in Gujarat and India.

### Questionnaires

Two questionnaires were developed: one in Gujarati, for those who could read and write in Gujarati, which asked about patients’ experiences and one in English, for those who could read and write in English, which asked about patients’ experiences. The questions were based on those used in an earlier Swedish study on Ideas, Concerns & Expectations ^6^. As the previously existing questionnaires included all the items wanted in this study, we based the questionnaire on the existing one instead of creating new one. A research team made up of experts in both the subject matter and language carefully reviewed, translated, and based the questions to ensure accuracy and appropriateness in both English and Gujarati. This involved forward–backward translation and expert review to ensure the meaning remained the same and that the terms related to ICE made sense and were relevant for the local people who would use it. The questions were about background information and ideas (Questions 2 and 3), concerns (Questions 5–8), expectations (Questions 9–11 and 13–14) and satisfaction (Questions 16–20) (Tables 2 and 3). Response options were ‘yes,’ ‘partly’, ‘no’ and ‘I don’t know’. Questions 1, 4, 12 and 15 were open-ended and were not included in this study but will be part of an upcoming qualitative study. The questionnaires were tested with 30 patients and three physiotherapists to ensure they were easy to understand and suitable for use by those who would be using them. After testing in a pilot study, no more changes were needed

### Sampling and ethics

Before starting the study, all managers and professionals who participated were fully informed about the study and agreed to participate voluntarily. The receptionists and researchers provided patients both spoken and written details about the study before they joined, explaining that participation was completely voluntary and that their identities would remain anonymous. The receptionists at the centres invited Gujarati-speaking adult patients to participate consecutively. They invited as many patients as possible during the study period who were booked for consultations with PTs and to keep track of the number of patients who agreed to participate. Only patients attending planned consultations were invited, not those attending acute care consultations. It was not possible to include acute care consultations because such consultations are too short for patients to provide reflections about their ICE.

After receiving written permission from the patients, the receptionist or researcher gave them a questionnaire in their language preferences. Immediately following the consultation, patients completed the questionnaires and returned them to the receptionist or researcher. The patients gave the completed questionnaire directly to the receptionist or researcher by physically handing it over. The receptionist retained the completed questionnaires until the researcher collected them. The participating centres varied in size and recruited different numbers of participants. The duration for which questionnaires were distributed and collected also varied by centre.

The study was approved by the Institutional Ethics Committee - CHARUSAT, Changa (Proposal Number: IEC/CHARUSAT/22/62).

### Data analysis

An initial investigation aimed at describing the patterns of patient ICEs and satisfaction in this specific context used descriptive statistics (frequencies and percentages). This approach helped identify prevalent patient perspectives and differences among various backgrounds and settings, thereby providing basic information necessary for future analytical studies examining relationships and mechanisms.

## Results

### Study population

Of 500 questionnaires given, 350 were filled out by patients and collected from the centres by one of the researchers. Data were analysed from 232 different patient consultations, involving 232 distinct patients who filled out a questionnaire. (Table 1). Sixty-eight patients who were asked to participate denied or returned the questionnaire (who consulted PTs). Most respondents were female. The majority of patients who consulted were 20-49 years old. The study’s main finding was that patient perspectives must be considered to reach an agreement during physiotherapy consultations. Patients’ opinions on the consultations differed, as shown by the study findings.

**Table 1.** Sex, age distribution and number of participants in 232 consultations with physiotherapists (PTs).

| Sex | Female | Male | Total |
| --- | --- | --- | --- |
| Age (years) | (127) | (105) | (232) |
| 20-49 | 69 | 57 | 126 |
| 50-69 | 51 | 41 | 92 |
| ≥70 | 7 | 7 | 14 |

Most patients who consulted physical therapists (PTs) experienced musculoskeletal problems.

This study analysed data at group level to identify patterns in patient responses.

### ICE responses

In consultations, 88.3% of patients said they felt comfortable talking about and explaining their problems. (Table 2, question 2). Approximately 89% of patients expressed that their questions about their health were answered by their healthcare providers (question 3). In 41.8% of consultations, patients reported sharing their worries (question 7). The number of visits to private physiotherapy centres was higher than that to centres managed by trusts (Table 4). 76% of patients who sought the advice of PTs expected to receive an explanation for their bodily sufferings.

**Table 2.** Question number and content, and distribution in percentages of answers from 232 patient consultations with physiotherapists (PT) regarding related ideas, concerns and expectations.

|  | % |  |  |  |
| --- | --- | --- | --- | --- |
| Areas | Yes | Partly yes | No | D/U |
| 2. If patients' thoughts and explanations about their symptoms emerged during the consultation. | 88.7% | 7.8% | 1.7% | 2.2% |
| 3. If patients' questions about health were answered | 89.6% | 8.2% | 0.9% | 1.7% |
| 5. If patients thought something was frightening that might cause their symptoms | 19.5% | 5.2% | 72.7% | 3.0% |
| 6. If patients had concerns and anxiety regarding investigation/treatment | 9.9% | 4.3% | 84.1% | 1.7% |
| 7. If patients' concerns were presented | 41.8% | 5.6% | 51.3% | 1.3% |
| 8. If patients had other concerns | 34.9% | 4.7% | 58.2% | 2.2% |
| 9. If patients had any expectations about receiving a reason/an explanation for their symptoms | 76.7% | 4.3% | 15.9% | 3.0% |
| 10. If patients felt they were respected and taken seriously | 87.9% | 3.9% | 6.9% | 1.3% |
| 11. If patients' expectations for the consultation were fulfilled | 90.9% | 6.9% | 0.4% | 1.7% |
| 13. If patients received help with what they expected | 92.2% | 6.5% | 0.9% | 0.4% |
| 14. If patients thought there was anything that should have been done at the consultation but was missed | 8.6% | 0.9% | 88.4% | 2.2% |

**Table 3.** Question number and content, and distribution in percentages of answers from 232 patient consultations with physiotherapists (PT) related satisfaction.

|  | % |  |  |  |
| --- | --- | --- | --- | --- |
| Areas | Yes | Partly yes | Partly no/No | D/U |
| 16. If patients were satisfied with how they behaved toward the consultation | 88.4% | 9.5% | 0.9% | 1.3% |
| 17. If patients felt they received enough information | 90.1% | 6.9% | 2.2% | 0.9% |
| 18. If patients felt involved in decisions about investigations and treatment | 91.4% | 6.0% | 1.7% | 0.9% |
| 19. If patients felt they received sufficient emotional support during the consultation | 94.4% | 3.0% | 1.7% | 0.9% |
| 20 If patients were satisfied with the consultations as whole | 93.1% | 5.2% | 0.4% | 1.3% |

**Table 4.** Location-wise Distribution in percentages from 232 patient consultations with physiotherapists (PTs) regarding the presentation of patient concerns.

| 7. If patient' concerns were presented |  |  | By Location |  |  |  |  |  |
| --- | --- | --- | --- | --- | --- | --- | --- | --- |
|  | Overall |  | Government |  | Private |  | Trust Managed |  |
| Options/Scal es | Frequency | % | Frequency | % | Frequency | % | Frequency | % |
| Yes | 97 | 42 | 24 | 43 | 45 | 42 | 28 | 41 |
| Partly yes | 13 | 6 | 2 | 4 | 8 | 7 | 3 | 4 |
| No | 119 | 51 | 29 | 52 | 53 | 49 | 37 | 54 |
| D/U | 3 | 1 | 1 | 2 | 2 | 2 | 0 | 0 |
| Total | 232 | 100 | 56 | 100 | 108 | 100 | 68 | 100 |

**Table 5.** Gender-wise distribution in percentages from 232 patient consultations with physiotherapists (PTs) regarding the presentation of patient concerns.

| 7. If patient' concerns were presented |  |  | By Gender |  |  |  |
| --- | --- | --- | --- | --- | --- | --- |
| Options/Scales | Overall |  | Male |  | Female |  |
|  | Frequency | % | Frequency | % | Frequency | % |
| Yes | 97 | 42% | 38 | 36% | 59 | 46% |
| Partly yes | 13 | 6% | 2 | 2% | 11 | 9% |
| No | 119 | 51% | 63 | 60% | 56 | 44% |
| D/U | 3 | 1% | 2 | 2% | 1 | 1% |
| Total | 232 | 100 | 105 | 100 | 127 | 100 |

### Patients’ satisfaction

87.9% (question 10) and 90.9% of patients (Table 2, question 11) felt that the consultation met their expectations. A few patients (question 14) said that something that should have been done was missing during their session. Patients (88.4–94.4%, questions 16–19) were satisfied with the PT’s treatment, the information and emotional support they received, and SDM. PT consultations at centres managed by trusts recorded the highest patient satisfaction rates (Table 8). Clients at various government physiotherapy centres reported feeling that they had received sufficient emotional support while consulting physiotherapists (Table 6).

**Table 6.** Location-wise distribution in percentages from 232 patient consultations with physiotherapists (PTs) to provide sufficient emotional support during consultation.

| 7. If patient' concerns were presented |  |  | By Gender |  |  |  |
| --- | --- | --- | --- | --- | --- | --- |
| Options/Scales | Overall |  | Male |  | Female |  |
|  | Frequency | % | Frequency | % | Frequency | % |
| Yes | 97 | 42% | 38 | 36% | 59 | 46% |
| Partly yes | 13 | 6% | 2 | 2% | 11 | 9% |
| No | 119 | 51% | 63 | 60% | 56 | 44% |
| D/U | 3 | 1% | 2 | 2% | 1 | 1% |
| Total | 232 | 100 | 105 | 100 | 127 | 100 |

**Table 7.**
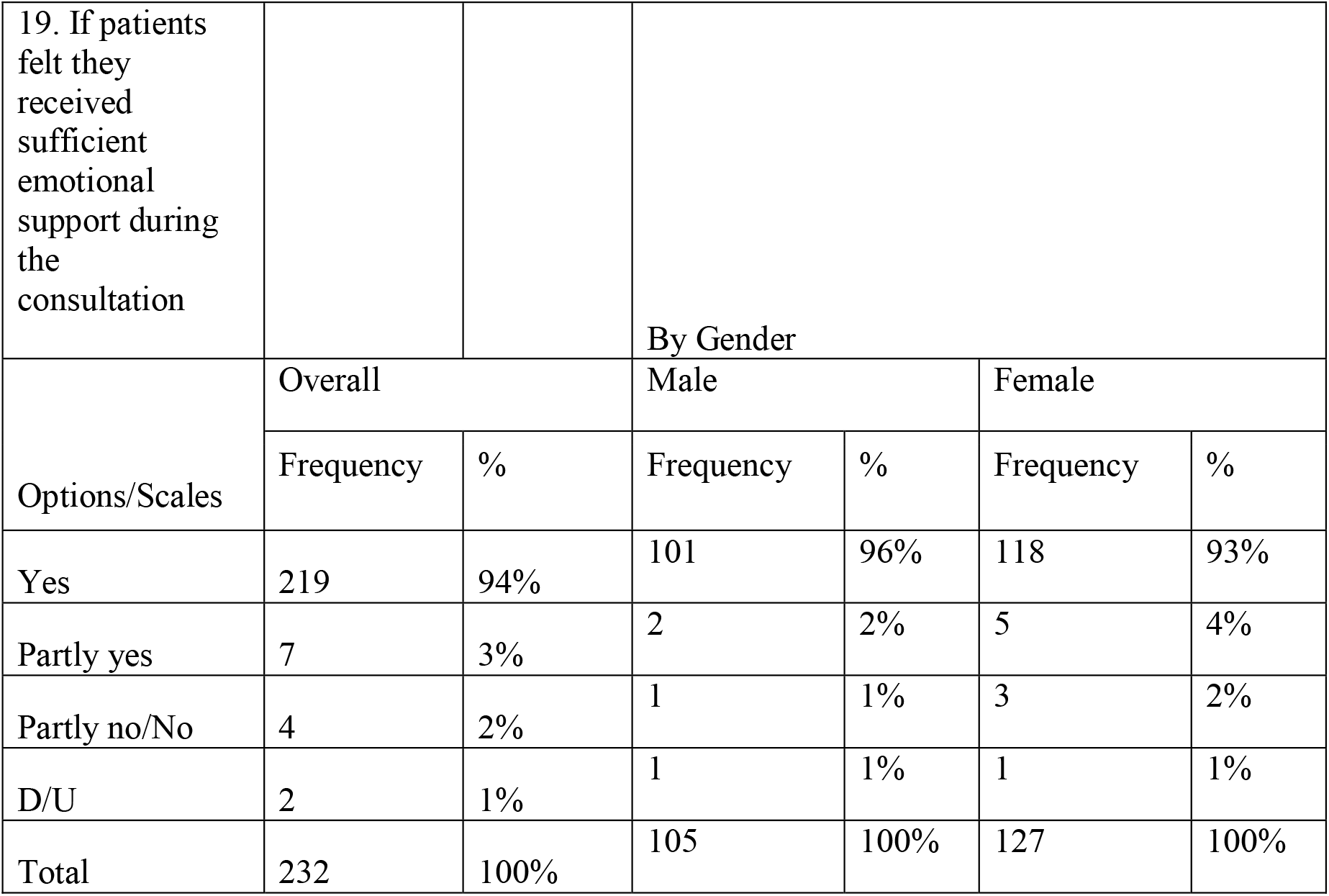
Gender-wise distribution in percentages from 232 patient consultations with physiotherapists (PTs) regarding receiving sufficient emotional support during the consultation.

|  |  |  |  |  |  |  |
| --- | --- | --- | --- | --- | --- | --- |
| 19. If patients felt they received sufficient emotional support during the consultation |  |  | By Gender |  |  |  |
| Options/Scales | Overall |  | Male |  | Female |  |
|  | Frequency | % | Frequency | % | Frequency | % |
| Yes | 219 | 94% | 101 | 96% | 118 | 93% |
| Partly yes | 7 | 3% | 2 | 2% | 5 | 4% |
| Partly no/No | 4 | 2% | 1 | 1% | 3 | 2% |
| D/U | 2 | 1% | 1 | 1% | 1 | 1% |
| Total | 232 | 100% | 105 | 100% | 127 | 100% |

**Table 8.** Location-wise distribution in percentages from 232 patient consultations with physiotherapists (PT) related satisfaction.

|  |  |  |  |  |  |  |  |  |
| --- | --- | --- | --- | --- | --- | --- | --- | --- |
| 20 If patients were satisfied with the consultations a whole |  |  | By Location |  |  |  |  |  |
| Options/Scales | Overall |  | Government |  | Private |  | Trust Managed |  |
|  | Frequency | % | Frequency | % | Frequency | % | Frequency | % |
| Yes | 216 | 93 | 54 | 96 | 95 | 88 | 67 | 99 |
| Partly yes | 12 | 5 | 1 | 2 | 11 | 10 | 0 | 0 |
| Partly no/No | 1 | 0 | 0 | 0 | 1 | 1 | 0 | 0 |
| D/U | 3 | 1 | 1 | 2 | 1 | 1 | 1 | 1 |
| Total | 232 | 100 | 56 | 100 | 108 | 100 | 68 | 100 |

**Table 9.** Gender-wise distribution in percentages from 232 patient consultations with physiotherapists (PT) related satisfaction.

|  |  |  |  |  |  |  |
| --- | --- | --- | --- | --- | --- | --- |
| 20 If patients were satisfied with the consultations a whole |  |  | By Gender |  |  |  |
|  | Overall |  | Male |  | Female |  |
| Options/Scales | Frequency | % | Frequency | % | Frequency | % |
| Yes | 216 | 93% | 101 | 96% | 115 | 91% |
| Partly yes | 12 | 5% | 2 | 2% | 10 | 8% |
| Partly no/No | 1 | 0% | 1 | 1% | 0 | 0% |
| D/U | 3 | 1% | 1 | 1% | 2 | 2% |
| Total | 232 | 100 | 105 | 100 | 127 | 100 |

## Discussion

This study offers patient perspectives on patient-centred communication practices within public and private physiotherapy clinics in a musculoskeletal setting in Gujarat, India. The study was conducted in a musculoskeletal setting.

HCPs and patients need to reach a shared understanding to improve how patients feel and encourage self-care management. The study revealed that most patients (88.3%) felt comfortable sharing their thoughts and ideas, however, fewer patients (41.8%) spoke up about their worries or concerns and despite this, many patients (76%) expected explanations for their symptoms. This difference, that patients find easier to express ideas but less so concerns (88.3% vs. 41.8%), shows that even though the open communication with patients, more sensitive issues might not be talked about. Previous studies have also found it difficult to get patients to talk about their deeper concerns, possibly because these concerns are sensitive or patients perceived communication barriers. This suggests that even if people generally feel satisfied overall, there may still be important concerns. This issue may be due to that it is sometimes difficult to discuss the most personal issue that a patient wants to talk about and this difficulty can be due to ineffective communication also. Neighbour and Larsen, found that it is important to build trust with their patients as it helps patients feel comfortable sharing unusual, personal, or embarrassing thoughts ^7 8^. Participants in this study visited their HCPs for follow-up visits or consultations. The appointments mentioned could have been scheduled either for routine health check-ups or for new health problems

Although the study found high satisfaction in participants, the results should be interpreted with caution as the questionnaire was filled out immediately after the consultation, which might have introduced social desirability bias. The satisfaction levels might be reported as higher than they really are suggesting that actual satisfaction levels or dissatisfaction might be hidden. Since 88.3% of participants said they felt comfortable sharing their ideas, suggests that patients felt heard and involved, likely an important factor in satisfaction. Further research is needed to understand this high satisfaction, especially since fewer expressions of concerns were reported by the participants ^9^.

According to the authors’ program theory, HCPs are more likely to involve patients in SDM when decisions are more difficult. However, earlier studies have shown that many geriatrics do not participate in SDM ^10,11^. Contrarily, patients in this study felt generally fairly involved in SDM. The participants’ high levels of education could explain the observed results ^12,13^. Patients with higher levels of education tend to take a more active role in shared decision-making (SDM) about their healthcare.

In the SDM process, previous relationships/connections help build trust which is important for the process. Trust influences choices for SDM, according to the results of other studies on older patients ^14^. The physiotherapy centre had a consistent group of staff members, so patients were likely to meet the same HCPs, helping their participation in SDM.

All HCPs involved in patient care participate in the SDM process to improve coordination and prevent gaps in care by helping to remove barriers between various health professionals ^15^. The “Ariadne principle” is a helpful method for consultation with patients who need to make shared decisions about their healthcare ^16^. The method affirms the need for patients to regularly review their priorities because of changing illnesses, symptoms as well as other life events that may change their priorities (such as the illness of their spouse or partner). Using a three-part consultation model leads to more complete and careful patient care ^17^. It better enables the ability to self-manage new symptoms ^18^. HCPs in India spend less time with their patients than HCPs in other countries ^19.^ Because India lacks enough doctors, it is difficult to get medical care, which causes more problems to arise at each visit.

Bodegård et al. found that patients who asked more questions during their medical visits felt more interrupted than patients who asked fewer questions ^20^. When a doctor carefully listens to a patient, the patient is less likely to feel dissatisfied. CCM treatments focus on encouraging patients to be knowledgeable and actively involved in their own healthcare to achieve better health outcomes ^21,22^.

The early idea of the SDM concept did not include goal setting ^23,24^. In the collaborative care model (CCM), it is important to set goals to aid self-management ^25^. Patients spoke more about ADL goals (such as being able to work from home or spend time with their children) than disease-related goals, which HCPs considered. Goals have seldom been discussed in home care programs when treating the elderly ^25–27^. Goal setting may not be given enough importance because HCPs find it uninteresting from HCPs’ perceptions and provide similarity for all clients ^27^.

The study included questions on satisfaction with consultations. Most patients were satisfied with their consultations. The study investigated whether factors other than consultation might affect satisfaction. In a cross-sectional study, patients with happy doctors reported better communication, consistency, access, and thoroughness ^17^. Factors, such as short wait times and reliable care, or patient characteristics (such as age and functional status), might influence satisfaction ^28–30^. It might be difficult to determine of the patients with the consultation itself could be hard to know.

Although the patients were highly satisfied, satisfaction alone does not directly imply treatment adherence in physiotherapy. Treatment adherence depends on many different factors beyond satisfaction, like delivery modality, clinician following to guidelines, cultural differences, and practical barriers ^31–33^. Although, this study found high satisfaction, it does not necessarily mean high adherence; future research should include explicit adherence measures.

### Strengths and weaknesses

This study explored patient perspectives. One of the strengths was a relatively large number of respondents. Because the study included several different centres, the results are likely generalisable. All centres involved were located in a higher socioeconomic status part of Gujarat compared to the rest of India, which negatively affects the generalisability of our findings, especially the high satisfaction levels reported, to other socioeconomic populations. Participants who took the questionnaire might not fully understand complex concepts such as ‘ideas,’ ‘concerns,’ and ‘expectations’, which can be a significant limitation. Different interpretations could have changed responses, therefore, the accuracy and detail of the reported ICE percentages may be questionable. An analysis of correlation between the patient agenda (ICE) and their satisfaction with the consultation was not conducted as overall patient satisfaction was good.

The immediate post-consultation filling of questionnaires may have limited patients to carefully consider their responses resulting in more superficial responses, especially regarding complex concerns or long-term satisfaction. Another limitation is that it is unclear whether this was the patient first consultation with an HCP at all, it was first time they consulted a PHC professional about this particular illness. It is possible that the behaviour of the professionals may have been influenced because they were aware that they were part of a research study. Analytical statistics were not used to present the findings because of the exploratory study design. A power calculation was not performed because the study was descriptive and the distribution of the collected data was unknown.

## Conclusion

This study investigates how patients and HCPs work together and how their ideas, concerns, and expectations (ICE) interact in a setting focused on musculoskeletal issues, and provides basic information that helps fill the gaps mentioned at the start of the paper about the specific context and population. The study also offers initial data that can be used for future research to better understand how particular parts of ICE work in this setting. A trustworthy PT–patient relationship supports patient-centred communication. A research study should be conducted to identify HCPs and family caregivers perceptions. The inclusion of communication with patients at core and SDM in the curriculum and its impact would be studied to improve HCP practice. Future studies could also investigate ICE and satisfaction in acute consultations to determine whether patterns are similar and explore HCP satisfaction with consultations.

## Data Availability

All data produced in the present study are available upon reasonable request to the authors

## Acknowledgements

The authors thank the government physiotherapy centres, private physiotherapy centres, trust centres with physiotherapy facilities, and everyone who participated in the study. The authors are also grateful to Dr. Adhish Trivedi, Dr. Dharmesh Parekh, PhD and Dr. Dhawal Kathrotiya for their wishes for the successful completion of this project and Vyoma Mistry for useful inputs on the manuscript.

